# The prevalence of burnout, risk factors and job-related stressors in gastroenterologists: a systematic review

**DOI:** 10.1101/2020.12.24.20248839

**Authors:** John Ong, Carla Swift, Michael Bath, Sharon Ong, Wanyen Lim, Yasseen Al-Naeeb, Arun Shankar, Yock Young Dan

**Affiliations:** Department of Engineering, University of Cambridge, Cambridge, UK; Department of Medicine, National University of Singapore, Singapore; Department of Gastroenterology, Norfolk & Norwich University Hospital NHS Trust, Norwich, UK; Department of Surgery, Whipps Cross University Hospital, Barts Health NHS Trust, London, UK; Division of Anaesthesiology and Perioperative Sciences, Singapore General Hospital, Singapore; Division of Anaesthesiology and Surgical Intensive Care, Sengkang General Hospital, Singapore; Department of Gastroenterology & Hepatology, Bedford Hospital NHS Trust, Bedford, UK

**Keywords:** Burnout, stress, gastroenterology, gastroenterologists, endoscopy

## Abstract

**Background and aims:** Clinician burnout is an important occupational hazard and the scale of the problem within gastroenterology remains poorly understood. The primary objective of this study was to identify the prevalence of burnout and its symptoms in gastroenterologists. The secondary objective was to identify risk factors and job-related stressors that commonly contribute to burnout in gastroenterologists.

**Methods:** Systematic searches were conducted in PubMed, Scopus, Cochrane and PsycINFO by two reviewers independently for articles published to 1 September 2020. The primary outcome measure was the reported prevalence of burnout in gastroenterologists. The secondary outcome measures were (i) the prevalence of burnout symptoms (emotional exhaustion, depersonalisation and low personal accomplishment) and (ii) the frequency of risk factors and stressors reported in studies. Data were tabulated and meta-analyses were presented as Funnel and Forest plots.

**Results:** Data were extracted from 11 studies. 54.5% (6/11) of these studies reported the prevalence of burnout in gastroenterologists; this ranged from 18.3% to 64.4%. Similar to burnout prevalence, burnout symptoms showed geographical variation and were common in gastroenterologists (up to 63.9%). Factors associated with work volume, age, and female gender were the three most frequently reported risk factors for increased levels of anxiety, stress or burnout in 72.7% (8/11), 54.5% (6/11), and 45.5% (5/11) of studies respectively. Significant methodological and clinical heterogeneity was observed.

**Conclusions:** Burnout and its symptoms are common in gastroenterologists but the syndrome is understudied within the field. Further research and good quality data are needed to help address the problem.

**Disclose Statement:** JO is funded by the British Society of Gastroenterology to conduct burnout research in the UK, and the Journal of Gastroenterology and Hepatology Foundation (Australia) to conduct burnout research in Southeast Asia.

## INTRODUCTION

In 2019, the World Health Organisation (WHO) recognised burnout as an ‘occupational phenomenon’ in the 11th revision of the International Classification of Disease (ICD).(1) The syndrome is characterised by abnormal feelings of emotional exhaustion (EE), depersonalisation (DP), and a sense of reduced personal accomplishment (PA). The latter two dimensions are also known as ‘cynicism’ and ‘professional efficacy’ respectively.(2) Symptoms exist on a scale of varying severity and affected individuals can go unrecognized due to the lack of awareness.(3) Importance is given to the early detection and management of burnout because troubling associations have been demonstrated between physician burnout, sub-optimal patient care, and physician ill health.(4–6) As such, it is an occupational hazard which requires well-placed support mechanisms to intervene, alleviate, or reverse burnout, ensuring the sustainability of the current workforce and the quality of care it delivers.(7,8)

In essence, burnout is precipitated by chronic and excessive stress at work. To that end, gastroenterology often involves long working hours, heavy workloads, large volumes of patients, invasive procedures and high amounts of stress. Furthermore, the current coronavirus disease 2019 (COVID-19) pandemic may have also compounded this risk, with early reports from severely affected countries suggesting that gastroenterologists have been placed under significantly higher amounts of stress due to the outbreak.(9– 11) Therefore, the scale of this problem must first be understood within gastroenterology in order to compel institutional improvements.(12)

The prevalence of burnout among gastroenterologists is currently unknown. Therefore, to understand the scale of burnout within the field, we conducted a PROSPERO registered systematic review of observational studies and randomised controlled trials on burnout. The primary objective of the systematic review was to understand the prevalence of burnout within gastroenterology. The secondary objectives were to: (i) understand the prevalence of burnout symptoms, and (ii) the risk factors and stressors that are commonly reported in gastroenterologists.

## METHODS

The methodology herein has been reported according to the PRISMA recommendations.(13) The protocol for this systematic review has been registered with PROSPERO, University of York (https://www.crd.york.ac.uk/prospero/), identification number CRD42020192707.

### Search strategy

Two authors, JO and CS, conducted systematic searches independently in PubMed, Scopus, PsycINFO and Cochrane databases from inception to 1st September 2020. Briefly, over 120 searches were conducted using search terms related, but not limited to, burnout, stress, gastroenterology, hepatology and endoscopy. The detailed search strategy is attached as Appendix 1 (Supplementary information). All articles were screened by title and abstract. Observational studies and randomised control trials that reported the (i) prevalence of burnout, (ii) prevalence of its symptoms, and (iii) stressors or risk factors associated with burnout were included for further eligibility screening. Our exclusion criteria were: (i) articles that did not report detailed quantitative data such as burnout prevalence rates, dimensional scores etc.; (ii) non-research articles e.g commentaries, editorials, correspondences etc; (iii) articles that did not report burnout data that is specific to gastroenterologists; (iv) low-quality observational studies defined by a modified Quality Assessment Tool for Systematic reviews of Observational studies (modified QATSO) score ≤ 33%(14), see Appendix 2 (Supplementary information); (v) articles that were not available through the UK Access Management Federation; and (vi) articles with irrelevant content. Any disagreements between JO and CS were resolved by consensus with WL and SO.

### Data extraction

All data were extracted into pre-designed forms on Microsoft Excel 2007 (Microsoft Corporation, Washington, US). Forms contained columns tabulating the country of origin of selected studies, first author including the year of publication, study population, sample size, response rates, reported prevalence rates of burnout, criteria for burnout, prevalence rates of EE, criteria for abnormal EE, prevalence rates of DP, criteria for abnormal DP, prevalence rates of PA, criteria for abnormal PA, stressors reported, and any risk factors associated with burnout. Risk factors and stressors were grouped under themes after a focus group discussion between JO, CS, WL and SO. Collated data were then summarised and presented as tables illustrated below.

### Statistical analyses

MedCalc 19.1.5 (MedCalc Software bv, www.medcalc.org) was utilised to perform the statistical analyses. Summary measures were the prevalence rates of burnout, the prevalence rates of EE, the prevalence rates of DP, and the prevalence rates of PA. Where applicable, meta-analyses were performed to obtain Funnel plots and Forest plots using reported prevalence rates (positive cases and sample sizes). I^2^ and Cochrane Q were used to test for heterogeneity between studies. Conventional classifications of the I^2^ statistic were used to describe heterogeneity; low, medium and high corresponded to I^2^ statistics of 25%, 50% and 75% respectively.(15) Random effect models were used to generate all Forest plots because medium to high heterogeneity was observed between studies. Egger’s test and Begg’s test were used to detect publication bias. Despite high heterogeneity between studies, meta-regression was not performed because the number of studies reporting burnout prevalence and its symptoms were less than 10 in both groups.(16) Data were therefore collated and presented in tables for better accuracy instead of pooled results.

## RESULTS

### Study selection and characteristics

A total of 808 articles were identified in the systematic search (Figure 1). After removing 490 duplicate search results, 318 articles were screened by title and abstracts. Of these, 237 articles had irrelevant content, 27 articles studied burnout in non- gastroenterology populations, and 39 articles were editorials, commentaries, letters or reviews. These were removed leaving 15 articles in the field of gastroenterology that were fully assessed for eligibility.(17–31) Subsequently, 4 articles were excluded from further analyses; 1 article had data which lacked sufficient quality(17) (modified QATSO score ≤ 33%) and 3 articles reported data on gastroenterology nurses only(18– 20). Overall, 11 studies were included in the qualitative synthesis.(21–31)

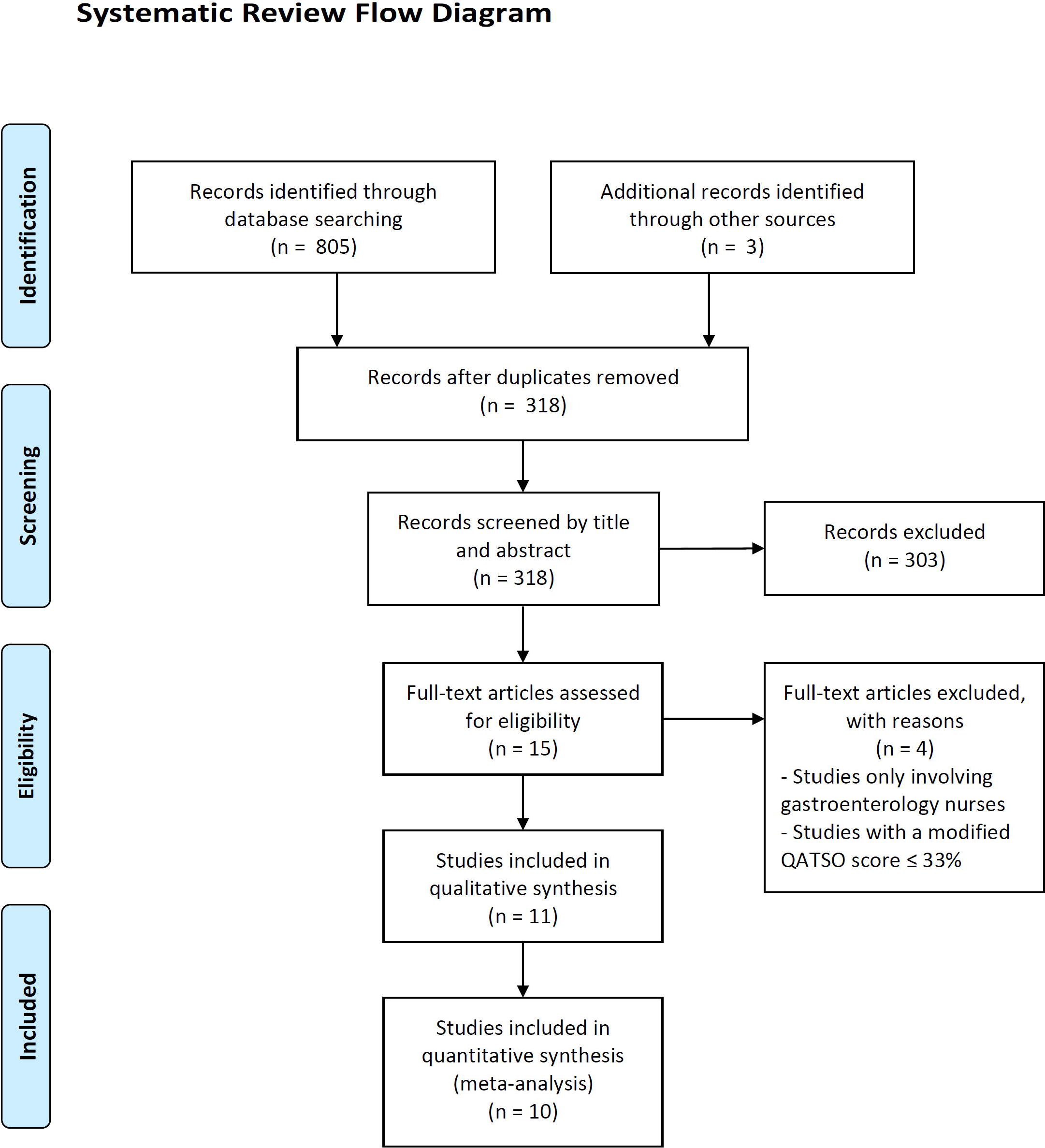

### Prevalence of burnout in gastroenterologists

54.5% (6/11) of studies reported a burnout prevalence in gastroenterologists. The median prevalence was 35.3% however the reported prevalence ranged broadly from 18.3% to 64.4%(21–26), see Table 1. Significant (“high”) heterogeneity was detected between the studies; I^2^ = 98.0% (95% CI: 97.2% to 98.6%) and Supplementary Figure 1 (Supplementary Information). No evidence of publication bias was detected (Egger’s test: p-value = 0.88). Subgroup analysis determined that methodological and clinical heterogeneity were present (Table 2). Respectively, in the six studies, there were three different burnout tools used to detect burnout in gastroenterologists and several different criteria were used to define burnout even when the same burnout detection tool was used. This, along with differences in study subpopulations (Table 2), contributed to a large degree of the heterogeneity observed in reported results.

**Table 1:** Reported prevalence of burnout in gastroenterologists. † Adjusted response rate = Number of gastroenterologists who had completed the burnout detection tool fully divided by the total target population. NR: not reported by the study. ‡ Additional data provided by authors through personal correspondence.

| Country | Authors | Sample Size | Adjusted Response Rate † | Burnout Prevalence |
| --- | --- | --- | --- | --- |
| Mexico | Aguilar-Nájera et al, 2020 (21) | 317 | 17.7%<br>(317/1792) | 27.8%<br>(88/317) |
| Mexico | Aguilar-Nájera et al, 2020 (21) | 301 | 22.0%<br>(301/1792) | 33.9%<br>(102/301) |
| International | Pawlak et al, 2020 (22) ‡ | 695 | NR | 18.3%<br>(122/666) |
| South Korea | Jang et al, 2019 (23) | 222 | NR | 64.4%<br>(143/222) |
| United Kingdom | Ong et al, 2020 (24) | 40 | 37.3%<br>(34/91) | 35.3%<br>(12/34) |
| United States & Canada | DeCross, 2017 (25) ‡ | 639 | 11.7%<br>(639/5440) | 54.3%<br>(347/639) |
| United States | Burke, 2017 (26) | 749 | 6.8%<br>(749/11080) | 49.0%<br>(367/749) |

**Table 2:** The frequency of burnout symptoms in gastroenterologists. 22-item MBI = Maslach Burnout Inventory Human Services Survey.(32) SIBOQ = Single Item Burnout Question.(33) § Single-item measure developed during the same study.

| Country | Authors | Study population | Burnout Detection Tool | Criteria for burnout | Burnout Prevalence Readout |
| --- | --- | --- | --- | --- | --- |
| Mexico | Aguilar-Nájera et al, 2020 (21) | Gastroenterologists, gastroenterology residents, endoscopists, endoscopy residents, gastroenterology fellows | 22-item MBI | EE > 27<br>OR<br>DP > 10<br>OR<br>PA < 33 | 27.8%<br>(88/317) |
| Mexico | Aguilar-Nájera et al, 2020 (21) | Gastroenterologists, gastroenterology residents, endoscopists, endoscopy residents, gastroenterology fellows | SIBOQ | Options 3 to 5 | 33.9%<br>(102/301) |
| International | Pawlak et al, 2020 (22) | Gastroenterology Trainees | SIBOQ | Options 3 to 5 | 18.3%<br>(122/666) |
| South Korea | Jang et al, 2019 (23) | Gastroenterologists | 22-item MBI | EE ≥ 27<br>OR<br>DP ≥ 10 | 64.4%<br>(143/222) |
| United Kingdom | Ong et al, 2020 (24) | Gastroenterology Trainees | 22-item MBI | EE ≥ 27 + DP ≥ 13<br>OR<br>EE ≥ 27 AND PA ≤ 31 | 35.3%<br>(12/34) |
| United States & Canada | DeCross, 2017 (25) | Gastroenterologists | 22-item MBI AND Single-item measure § | EE > 27 + DP > 10 + PA < 33 AND "Agreed" / "Strongly agreed" | 54.3%<br>(347/639) |
| United States | Burke, 2017 (26) | Gastroenterologists | 22-item MBI | EE ≥ 27<br>OR<br>DP ≥ 13 | 49.0%<br>(367/749) |

### Symptoms of burnout in gastroenterologists

54.5% (6/11) of studies reported data on the symptoms of burnout in gastroenterologists.(21,23,24,27–29) Abnormal symptoms (EE, DP or PA) in gastroenterologists were reported in all study populations to varying degrees. These varied geographically even though there was less heterogeneity in definitions of abnormal cut-offs for EE, DP and PA values (Table 3) compared to the definitions used to identify burnout. Emotional exhaustion was present in gastroenterologists across all studies ranging from 7.4% to 57.5%, showing less heterogeneity in funnel plots compared to DP and PA; Supplementary Figures 2 - 4 (Supplementary Information). DP varied from 0% to 48.7% and PA varied from 1.5% to 63.9%.

**Table 3:** The frequency of burnout symptoms in gastroenterologists.

| Country | Author | Abnormal EE | Abnormal DP | Abnormal PA |
| --- | --- | --- | --- | --- |
| Germany | Adarkwah et al 2020 (27) | 16.6% (113/683)<br>Criterion: EE ≥ 27 | 27.7% (189/683)<br>Criterion: DP ≥ 10 | 53.9% (368/683)<br>Criterion: PA ≤ 33 |
| Mexico | Aguilar-Nájera et al, 2020 (21) | 20.4% (84/411)<br>Criterion: EE > 27 | 10.7% (44/411)<br>Criterion: DP > 10 | 3.6% (15/411)<br>Criterion: PA < 33 |
| South Korea | Jang et al, 2019 (23) | 53.2% (118/222)<br>Criterion: EE ≥ 27 | 48.7% (108/222)<br>Criterion: DP ≥ 10 | 52.3% (116/222)<br>Criterion: PA ≤ 33 |
| United States | Keswani et al, 2011 (28) | 7.4% - 11.1%<br>Criterion: NR | 0%<br>Criterion: NR | 1.5% - 3.7%<br>Criterion: NR |
| United Kingdom | Ong et al, 2020 (24) | 57.5% (23/40)<br>Criterion:<br>EE > mean + 0.5 SD* | 23.5% (8/34)<br>Criterion:<br>DP > mean + 1.25 SD* | 63.9% (23/36)<br>Criterion:<br>PA < mean + 0.1 SD* |
| United Kingdom | Ramirez et al, 1996 (29) | 31.0% (75/241)<br>Criterion: EE ≥ 27 | 28.0% (67/241)<br>Criterion: DP ≥ 10 | 38.0% (91/241)<br>Criterion: PA ≤ 33 |

### Stressors and risk factors for burnout in gastroenterologists

11 studies(21–31) reported stressors or risk factors for burnout in gastroenterologists. Factors related to high work volumes were the most reported risk factors associated with higher stress or more severe symptoms of burnout in 72.7% (8/11) of studies. Of the remaining 3 studies, 2 did not comment on factors related to work volume(22,27) and 1 reported no relationship between workload and burnout(31). Age was the second most reported risk factor; 54.5% (6/11) of studies. The reported ages for at-risk groups were heterogeneous: 40 to 49 years(30), age under 48 years(27), age 50.4 ± 9.2 years(26), and age ≤ 55 years(29). 2 studies defined age as “younger” without specifying numerical values.(28,31) Of the remaining 5 studies, 3 did not report any age-related comparisons(21,22,25) and 2 found no significant associations(23,24).

The third most reported risk factor was female gender. 45.5% (5/11) of studies reported that female gastroenterologists experienced more anxiety, stress or burnout than male gastroenterologists. In the remaining 6 studies, 2 studies reported no difference in burnout prevalence rates between genders(21,24), 3 studies reported lower PA scores in females(27,28,31) but did not report gender-specific burnout rates, and 1 study did not report any data on gender comparisons(29). Regardless of burnout tools or criteria of burnout, a meta-analysis of 2,694 gastroenterologists across all 6 studies that reported burnout prevalence suggested that female gastroenterologists may be at higher risk of burnout compared to their male counterparts; OR = 1.5 (95% CI: 1.1 to 2.1), p < 0.01 (Supplementary Figure 5). Lack of support from colleagues and performing endoscopic procedures were other important stressors reported in 27.3% (3/11) of studies. Table 4 provides a summary of the risk factors and job-related stressors in gastroenterologists.

**Table 4:** Stressors and risk factors for burnout in gastroenterologists.

| Themes | Stressors and risk factors associated with anxiety, stress, and burnout |
| --- | --- |
| **Individual factors** | Female gender (22,23,25,26,30)<br>Age (26–31)<br>Lack of confidence (30)<br>Medical knowledge required for normal practice (25) |
| **Factors related to work volume** | High workload (24,29,30)<br>Longer working hours (28)<br>Work-life imbalance (23,25,26)<br>Non-medical duties during work (21) |
| **Factors related to professional risk or responsibility** | Endoscopic procedures (21,25,28)<br>Clinical errors (30)<br>Managerial responsibilities (29) |
| **Factors related to resources at work** | Inadequate staffing levels (24)<br>Non-user friendly IT systems (26)<br>Poor management of resources (29)<br>Lack of resources (29)<br>Lack of Personal Protective Equipment during COVID-19 (22)<br>Working conditions beyond control (30) |
| **Factors related to training** | Length of training (22,24)<br>Postgraduate exams (24)<br>Lack of training in communication and management skills (29)<br>Lack of endoscopic training during COVID-19 (22)<br>Curriculum requirements (24) |
| **Factors related to relationships with colleagues** | Lack of support from colleagues (21,24,30)<br>Conflict / violence at work (21,30)<br>The expectation of seniors (24)<br>Reprimands by seniors (21)<br>Accusations of inappropriate behaviour (30) |
| **Factors related to the location of work** | A long commute to work (24)<br>Living in a big city (21) |
| **Factors related to patient relationships** | Conflict / violence at work (21,30) |
|  | Dealing with patient suffering (29) |
|  | Difficult patients and managing patient expectations (24) |
|  | Accusations of inappropriate behaviour (30) |
| **Others** | Lack of awareness or provision for mental health services (22,24) |
|  | The increasing imposition of external regulatory burdens (25) |
|  | Work unrelated to medicine (21) |

## DISCUSSION

Occupational burnout was first described in the 1970s (34,35) and research in the area remains popular in the field of medicine. Despite this and nearly fifty years on, our study has demonstrated that this phenomenon remains poorly studied within gastroenterology. To date, we have identified only 7 studies that report the prevalence of burnout within the speciality; 6 in gastroenterologists and 1 in gastroenterology nurses. Furthermore, significant methodological and clinical heterogeneity between these studies, and the lack of good quality data, limit direct comparisons and further in-depth meta-analyses.

Similar to systematic reviews of burnout prevalence in medicine and other specialities(36,37), the differences in burnout tools and criteria to identify burnout contributed significantly to methodological heterogeneity (Table 2). The 22-item MBI is the most commonly used and extensively validated tool to detect burnout in clinicians(36,38) although it does have limitations, such as cost to administer and survey length. Nonetheless, only 66.7% (4/6) of the studies that reported burnout prevalence in gastroenterologists used this tool. In the remaining 2 studies, a less reliable single-item measure of burnout was used. Although shorter surveys may be cost-free and improve response rates, it has been shown that abbreviated versions of the 22-item MBI and single-item measures of burnout can be inaccurate.(39,40) As such, in the absence of a universally accepted tool, some researchers continue to advise against the use of these tools because readouts may be unreliable.(12,39,41).

Another crucial factor that contributes to clinical heterogeneity in burnout prevalence is the criteria used to define burnout when the 22-item MBI is used. In the earlier editions of the MBI, abnormal scores in all three burnout components were needed to identify burnout (High EE AND high DP AND low PA scores). However, some researchers have adapted the criteria in various ways to improve burnout detection. Subsequently, the only criteria of burnout that was validated clinically against symptoms in the WHO ICD are “a high EE score AND a high DP score” or “a high EE score AND a low PA score”. This has been supported by Maslach and corroborated by other researchers.(40,42,43) Only 1 study that was identified used these criteria to define burnout. If the above criteria is used in the above-mentioned studies, then up to 16-20% of gastroenterologists in Mexico and Germany, 30-40% in the UK, and 50-55% in South Korea, Canada and the United States approximately, would either have burnout or be at high risk of developing burnout. Abnormal scores in one dimension alone should not be used to define the presence of burnout because it is conceptually inadequate and these can be present in individuals without burnout i.e. different personality or profile types.(32,44,45) Therefore, using single-dimension criteria confers a significant risk of overestimating burnout prevalence.

This systematic review has also demonstrated that a pooled prevalence of burnout is inappropriate because burnout symptoms varied significantly across studies despite near-uniform cut-off values for abnormal subcomponent (EE, DP and PA) scores. This heterogeneity may therefore reflect actual differences in burnout prevalence as a result of different healthcare systems, countries, cultures and the prevalence of mental health disease. Pooled prevalence by geographical region may be attempted in the future however more burnout studies are needed globally. Reassuringly, native-language versions of the MBI-HSS have been previously validated in the countries within Table 2 and so the MBI-HSS has the potential to accurately identify burnout in these populations in future studies, provided rigorous burnout criteria are used. It is also noteworthy that previous scores on the MBI scale were derived arbitrarily(32) and therefore delineations of “low”, “medium” and “high” subscale scores carry limited clinical value. Therefore, Maslach has recommended the use of a different method to identify abnormal symptom scores. This is referred to as “Method 2 (AVE)” in the fourth edition of the MBI manual.(32). Advantages are three-fold; firstly the computed score remains valid even if not all questions are completed by respondents, secondly, a normal range based on over six thousand US healthcare workers has been established for comparison, and thirdly normal ranges for different populations can be tailored if needed. Therefore, it becomes important for researchers to report results using both methods in the future to allow comparison of results across different times and populations.

In terms of stressors and risk factors for burnout, it is unsurprising that factors related to high work volumes were reported in the majority (72.7%) of studies since burnout is typically precipitated by work-related stress. However, it is unusual that gastroenterologists aged approximately between their late thirties and early fifties were reported to be more prone to anxiety, stress and burnout. Further research is needed to explore these observations. Interestingly, female gastroenterologists were reported to be at higher risk of suffering from anxiety, stress and burnout. The association between female gender and anxiety may be consistent with population-based studies(22,46) and having added domestic commitments may have contributed to stress in female gastroenterologists(23). Nonetheless, having organisation-led initiatives, such as mentoring schemes and easily accessible wellbeing support services, can help lower anxiety, stress and burnout risk.(17,22,47,48) Furthermore, regardless of age and gender, access to support services can also aid in the early detection and management of underlying mental health conditions which can also be associated with burnout.(49) Surprisingly, factors associated with relationships with colleagues (lack of support, conflict, high expectations etc) were a frequently reported source of stress for gastroenterologists. These stressors are modifiable and could potentially be addressed through measures such as improving communication and mindfulness-based activities, both of which have been reported to have demonstrable effects in large meta-analyses.(50,51)

This study had several limitations. Firstly, large scale burnout studies are invariably survey based and the response rates in the included studies were low. Therefore, survey bias such as response, non-response and sampling bias could not have been excluded. This could have affected the meta-analyses on burnout in female gastroenterologists although high heterogeneity and publication bias were not detected. Secondly, there were insufficient high-quality data for meta-analyses to determine the strengths of associations between stressors and other risk factors of burnout, e.g. age. Thirdly, all but one study were conducted before the COVID-19 outbreak therefore previously reported burnout rates may not be reflective of burnout rates during the pandemic.

## CONCLUSION

Burnout and its symptoms are common in gastroenterologists but the syndrome is understudied within the field. Further research and good quality data are needed to help address the problem.

## Data Availability

All data have been provided and available online.

## Acknowledgements

Communications and access to literature were provided by the University of Cambridge. JO is funded by the W.D. Armstrong Doctoral Research Training Fellowship at the University of Cambridge and a talent development grant from the National University of Singapore.

## Author’s contributions

Conceptualisation: JO; design: JO, MB, WL, YAN; data curation JO and CS; data analysis: all authors; manuscript preparation: JO, CS and MB; revisions: JO, CS, MB, WL, SO, YAN and AS; critical edits and supervision: YYD.

## Notes

### Competing Interest Statement

The authors have declared no competing interest.

### Clinical Protocols

https://www.crd.york.ac.uk/PROSPEROFILES/192707_PROTOCOL_20201020.pdf

### Author Declarations

This is a systematic review. Ethical approval was not needed.

