## Supplementary Files for "The prevalence of burnout, risk factors and job-related stressors in gastroenterologists: a systematic review"

#### Supplementary Information


| Contents | Item description | Page number |
|---|---|---|
| Appendix 1 | Systematic review search strategy | 2 to 4 |
| Appendix 2 | Modified QATSO tool | 5 |
| Supp. Figure 1 | Burnout prevalence: Forest & Funnel plots | 6 |
| Supp. Figure 2 | EE prevalence: Forest & Funnel plots | 7 |
| Supp. Figure 3 | DP prevalence: Forest & Funnel plots | 8 |
| Supp. Figure 4 | PA prevalence: Forest & Funnel plots | 9 |
| Supp. Figure 5 | Burnout in female gastroenterologists (OR) | 10 |

#### Appendix 1: Systematic review search strategy

"Term 1" AND "Term 2" (AND "Term 3" where indicated)

Period: From inception to 1 September 2020

| Search | Term 1 | Term 2 | Term 3 |
|---|---|---|---|
| 1 | Burn out | Gastroenterology | - |
| 2 | Burn out | Gastroenterologists | - |
| 3 | Burn out | Endoscopy | - |
| 4 | Burn out | Endoscopists | - |
| 5 | Burn out | Gastrointestinal | - |
| 6 | Burn out | GI | - |
| 7 | Burn out | Hepatology | - |
| 8 | Burn out | Hepatologists | - |
| 9 | Burnout | Gastroenterology | - |
| 10 | Burnout | Gastroenterologists | - |
| 11 | Burnout | Endoscopy | - |
| 12 | Burnout | Endoscopists | - |
| 13 | Burnout | Gastrointestinal | - |
| 14 | Burnout | GI | - |
| 15 | Burnout | Hepatology | - |
| 16 | Burnout | Hepatologists | - |
| 17 | Burned out | Gastroenterology | - |
| 18 | Burned out | Gastroenterologists | - |
| 19 | Burned out | Endoscopy | - |
| 20 | Burned out | Endoscopists | - |
| 21 | Burned out | Gastrointestinal | - |
| 22 | Burned out | GI | - |
| 23 | Burned out | Hepatology | - |
| 24 | Burned out | Hepatologists | - |
| 25 | Burnt out | Gastroenterology | - |
| 26 | Burnt out | Gastroenterologists | - |
| 27 | Burnt out | Endoscopy | - |
| 28 | Burnt out | Endoscopists | - |
| 29 | Burnt out | Gastrointestinal | - |
| 30 | Burnt out | GI | - |
| 31 | Burnt out | Hepatology | - |
| 32 | Burnt out | Hepatologists | - |
| 33 | Emotional exhaustion | Gastroenterology | - |
| 34 | Emotional exhaustion | Gastroenterologists | - |
| 35 | Emotional exhaustion | Endoscopy | - |
| 36 | Emotional exhaustion | Endoscopists | - |
| 37 | Emotional exhaustion | Gastrointestinal | - |

| | | | |
|---|---|---|---|
| 38 | Emotional exhaustion | GI | - |
| 39 | Emotional exhaustion | Hepatology | - |
| 40 | Emotional exhaustion | Hepatologists | - |
| 41 | Depersonalisation | Gastroenterology | - |
| 42 | Depersonalisation | Gastroenterologists | - |
| 43 | Depersonalisation | Endoscopy | - |
| 44 | Depersonalisation | Endoscopists | - |
| 45 | Depersonalisation | Gastrointestinal | - |
| 46 | Depersonalisation | GI | - |
| 47 | Depersonalisation | Hepatology | - |
| 48 | Depersonalisation | Hepatologists | - |
| 49 | Cynicism | Gastroenterology | - |
| 50 | Cynicism | Gastroenterologists | - |
| 51 | Cynicism | Endoscopy | - |
| 52 | Cynicism | Endoscopists | - |
| 53 | Cynicism | Gastrointestinal | - |
| 54 | Cynicism | GI | - |
| 55 | Cynicism | Hepatology | - |
| 56 | Cynicism | Hepatologists | - |
| 57 | Professional efficacy | Gastroenterology | - |
| 58 | Professional efficacy | Gastroenterologists | - |
| 59 | Professional efficacy | Endoscopy | - |
| 60 | Professional efficacy | Endoscopists | - |
| 61 | Professional efficacy | Gastrointestinal | - |
| 62 | Professional efficacy | GI | - |
| 63 | Professional efficacy | Hepatology | - |
| 64 | Professional efficacy | Hepatologists | - |
| 65 | Personal accomplishment | Gastroenterology | - |
| 66 | Personal accomplishment | Gastroenterologists | - |
| 67 | Personal accomplishment | Endoscopy | - |
| 68 | Personal accomplishment | Endoscopists | - |
| 69 | Personal accomplishment | Gastrointestinal | - |
| 70 | Personal accomplishment | GI | - |
| 71 | Personal accomplishment | Hepatology | - |
| 72 | Personal accomplishment | Hepatologists | - |
| 73 | Stress | Gastroenterology | Burn out |
| 74 | Stress | Gastroenterologists | Burn out |
| 75 | Stress | Endoscopy | Burn out |
| 76 | Stress | Endoscopists | Burn out |
| 77 | Stress | Gastrointestinal | Burn out |
| 78 | Stress | GI | Burn out |
| 79 | Stress | Hepatology | Burn out |
| 80 | Stress | Hepatologists | Burn out |
| 81 | Stress | Gastroenterology | Burnout |
| 82 | Stress | Gastroenterologists | Burnout |
| 83 | Stress | Endoscopy | Burnout |

| | | | |
|---|---|---|---|
| 84 | Stress | Endoscopists | Burnout |
| 85 | Stress | Gastrointestinal | Burnout |
| 86 | Stress | GI | Burnout |
| 87 | Stress | Hepatology | Burnout |
| 88 | Stress | Hepatologists | Burnout |
| 89 | Stress | Gastroenterology | Burned out |
| 90 | Stress | Gastroenterologists | Burned out |
| 91 | Stress | Endoscopy | Burned out |
| 92 | Stress | Endoscopists | Burned out |
| 93 | Stress | Gastrointestinal | Burned out |
| 94 | Stress | GI | Burned out |
| 95 | Stress | Hepatology | Burned out |
| 96 | Stress | Hepatologists | Burned out |
| 97 | Stressors | Gastroenterology | Burn out |
| 98 | Stressors | Gastroenterologists | Burn out |
| 99 | Stressors | Endoscopy | Burn out |
| 100 | Stressors | Endoscopists | Burn out |
| 101 | Stressors | Gastrointestinal | Burn out |
| 102 | Stressors | GI | Burn out |
| 103 | Stressors | Hepatology | Burn out |
| 104 | Stressors | Hepatologists | Burn out |
| 105 | Stressors | Gastroenterology | Burnout |
| 106 | Stressors | Gastroenterologists | Burnout |
| 107 | Stressors | Endoscopy | Burnout |
| 108 | Stressors | Endoscopists | Burnout |
| 109 | Stressors | Gastrointestinal | Burnout |
| 110 | Stressors | GI | Burnout |
| 111 | Stressors | Hepatology | Burnout |
| 112 | Stressors | Hepatologists | Burnout |
| 113 | Stressors | Gastroenterology | Burned out |
| 114 | Stressors | Gastroenterologists | Burned out |
| 115 | Stressors | Endoscopy | Burned out |
| 116 | Stressors | Endoscopists | Burned out |
| 117 | Stressors | Gastrointestinal | Burned out |
| 118 | Stressors | GI | Burned out |
| 119 | Stressors | Hepatology | Burned out |
| 120 | Stressors | Hepatologists | Burned out |

### Appendix 2: Modified QATSO Tool

| S/N | Criteria | Scores |
|---|---|---|
| 1 | Was the sampling method representative of the population intended to the study?<br>A. Non-probability sampling (including: purposive, quota , convenience and snowball sampling)<br>B. Probability sampling (including: simple random, systematic, stratified g, cluster, two-stage and multi-stage sampling)<br>. | <br>0<br><br>1 |
| 2 | Were the measurement of burnout, frequency of abnormal symptoms in the study population, or related stressors reported?<br>A. Burnout prevalence<br>B. Frequency of burnout symptoms<br>C. Stressors related burnout<br>D. None of the above reported | <br>1<br>1<br>1<br>0 |
| 3 | Did the study report any response rate? (If the reported response rate is below 60%, the question should be answered "No".)<br>A. No<br>B. Yes | <br>0<br>1 |
| 4 | Did the investigator(s) control for confounding factors (e.g. stratification/ matching/ restriction/ adjustment) when analyzing the associations (if the study contains purely descriptive results, no association and prediction tests were conducted in the test, please select "Not applicable")?<br>A. No<br>B. Yes<br>C. Not applicable | <br>0<br>1<br>N/A |

Scoring method: Total score divided by total number of all applicable items

Grading of the modified QATSO score:

| 0% -33% | 33%- 66% | 67% -100% |
|---|---|---|
| Poor | Satisfactory | Good |

Studies scoring 33% or lower (score ≤ 2) were excluded from meta-analysis.

**Supplementary Figure 1 - Meta-analyses of Burnout Prevalence in Gastroenterologists**

**(A)**

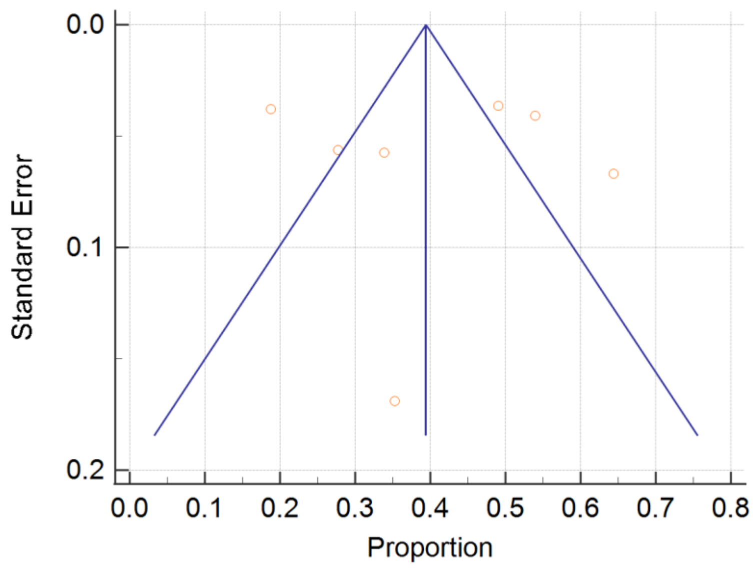


**(B)**

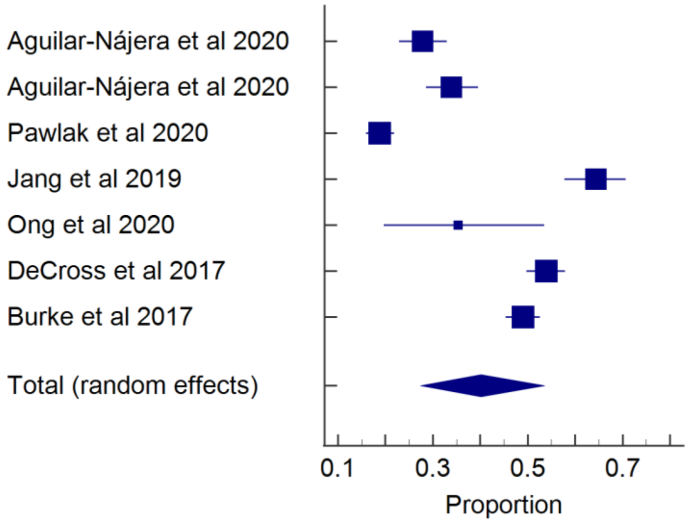


**Cochrane Q = 305.1, df = 6, p < 0.0001**

**$I^2$ = 98.0% (95% CI: 97.2% to 98.6%)**

**Pooled prevalence = 40.1% (95% CI: 27.4% to 53.6%)**

**Egger's test: p = 0.88, Begg's test: p = 0.65**

Supplementary Figure 1: (A) Funnel plot and (B) Forrest plot of studies reporting burnout prevalence in gastroenterologists.

**Supplementary Figure 2: Prevalence of emotional exhaustion (EE) in gastroenterologists.**

**(A)**

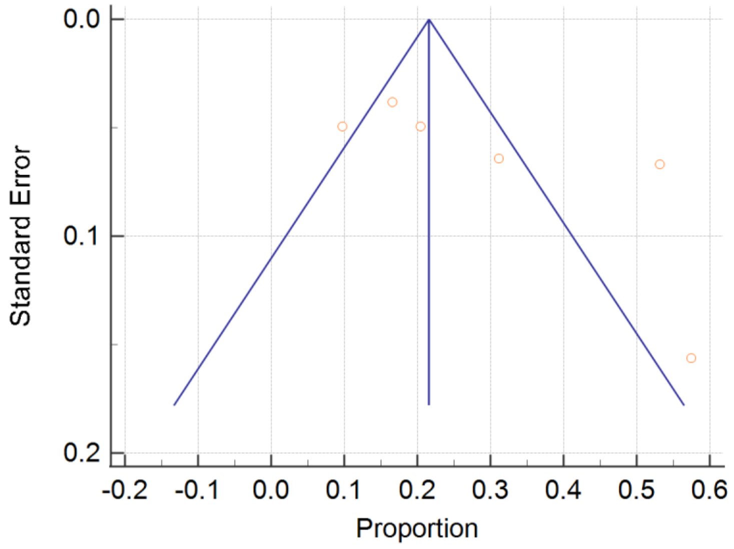


**(B)**

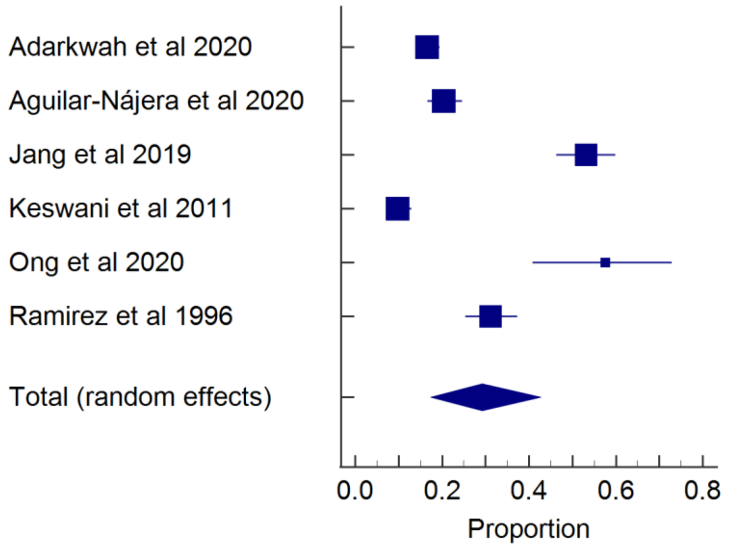


**Cochrane Q = 189.5, df = 5, p < 0.0001**

**$I^2$ = 97.4% (95% CI: 95.9% to 98.3%)**

**Pooled prevalence = 29.2% (95% CI: 17.4% to 42.7%)**

**Egger's test: p = 0.13, Begg's test: p = 0.09**

Supplementary Figure 2: (A) Funnel plot and (B) Forrest plot of studies reporting the prevalence of emotional exhaustion in gastroenterologists.

**Supplementary Figure 3 - Prevalence of depersonalisation (DP) in gastroenterologists.**

**(A)**

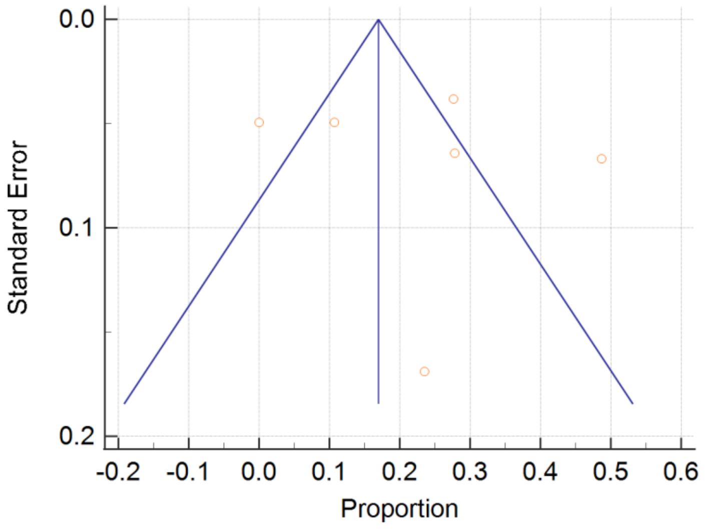


**(B)**

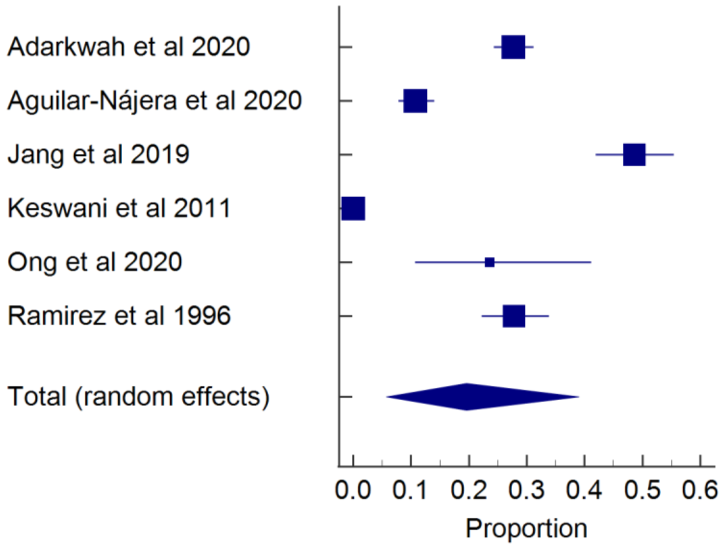


**Cochrane Q = 447.8, df = 5, p < 0.0001**

**$I^2$ = 98.9% (95% CI: 98.5% to 99.2%)**

**Pooled prevalence = 19.6% (95% CI: 5.7% to 39.0%)**

**Egger's test: p = 0.79, Begg's test: p = 0.85**

Supplementary Figure 3: (A) Funnel plot and (B) Forrest plot of studies reporting the prevalence of depersonalisation in gastroenterologists.

**Supplementary Figure 4 - Prevalence of low personal accomplishment (PA) in gastroenterologists.**

**(A)**

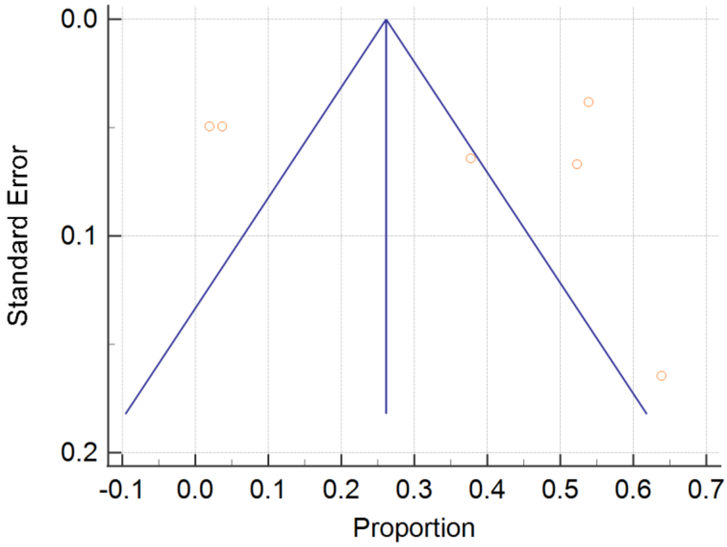


**(B)**

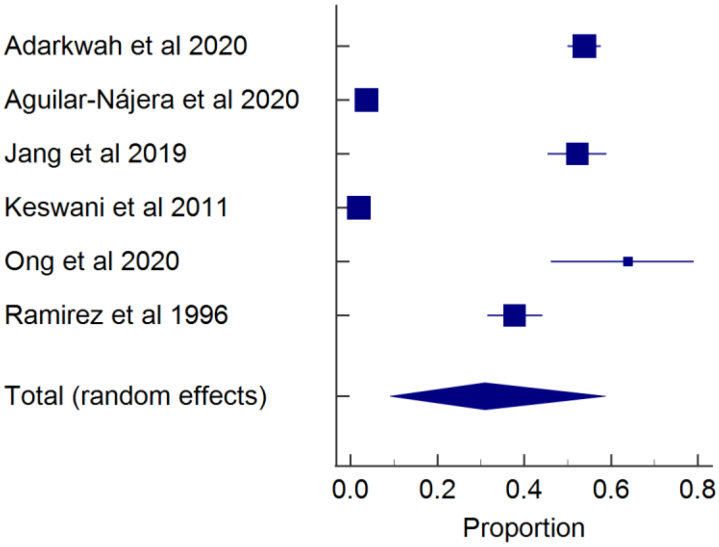


**Cochrane Q = 774.4, df = 5, p < 0.0001**

**$I^2$ = 99.4% (95% CI: 99.2% to 99.5%)**

**Pooled prevalence = 30.9% (95% CI: 9.1% to 58.8%)**

**Egger's test: p = 0.87, Begg's test: p = 0.07**

Supplementary Figure 4: (A) Funnel plot and (B) Forrest plot of studies reporting the prevalence of low personal accomplishment in gastroenterologists.

**Supplementary Figure 5 - Burnout in female gastroenterologists (Odd's Ratio)**

**(A)**

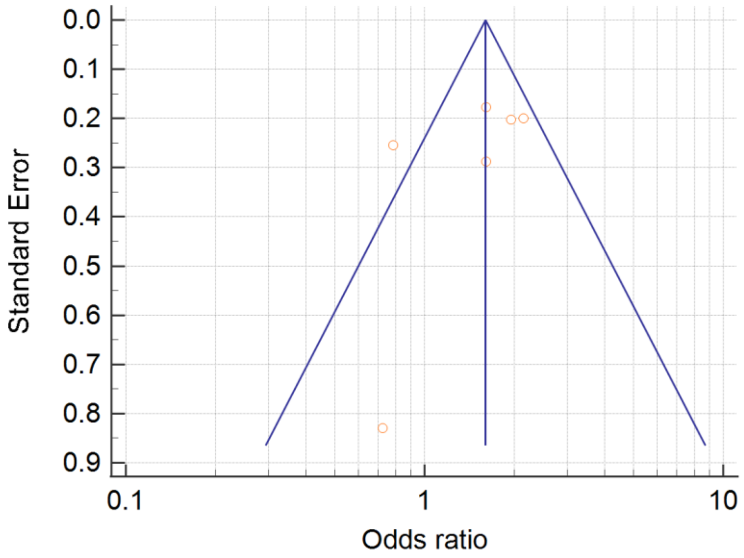


**(B)**

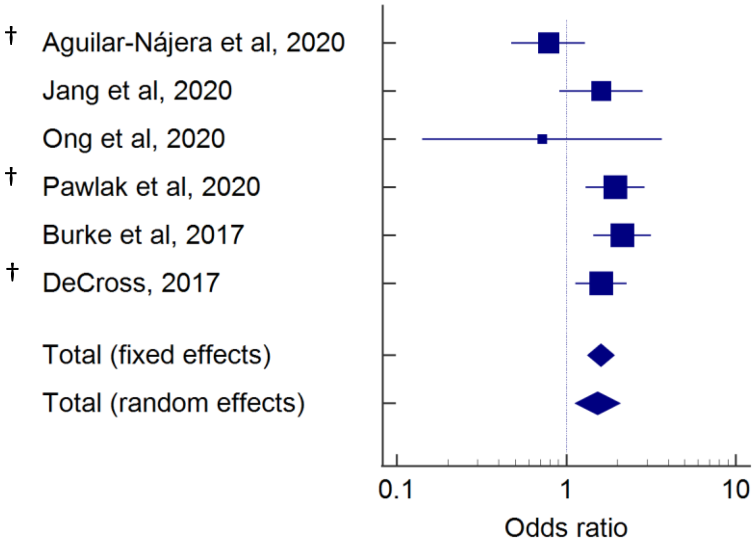


**Cochrane Q = 11.9, df = 5, p = 0.04**

**$I^2$ = 57.8% (95% CI: 0% to 82.9%)**

**Pooled Odd's Ratio (random effect) = 1.5 (95% CI: 1.1 to 2.1), p < 0.01**

**Egger's test: p = 0.36, Begg's test: p = 0.35**

Supplementary Figure 5: (A) Funnel plot and (B) Forrest plot showing Odd's Ratio of studies reporting gender-specific burnout rates in female gastroenterologists. †Additional data provided by authors through personal correspondence.
